# Sociodemographic correlates of parent and youth-reported eating disorder symptoms in the Adolescent Brain Cognitive Development Study

**DOI:** 10.1101/2023.12.18.23300155

**Authors:** Carolina Makowski, Margaret L. Westwater, Kyung E. Rhee, Jingjing Zou, Amanda Bischoff-Grethe, Christina E. Wierenga

## Abstract

**Purpose:** Eating Disorders (EDs) often start in adolescence, though ED-related concerns in diverse youth samples remain understudied. We leveraged data from the Adolescent Brain Cognitive Development□(ABCD) Study to identify the prevalence of parent- and youth-reported ED symptoms and their sociodemographic characteristics.

**Methods:** Data were drawn from baseline (ages 9-11 years, n=11,868) and 2-year follow-up (ages 11-14 years; n=10,908) from the ABCD Study. A tetrachoric factor analysis summarized clusters of ED symptoms, which were compared between parent and youth reports and across sociodemographic variables.

**Results:** Three factors emerged reflecting “weight distress”, “weight control”, and “binge eating” (prevalence range: 1.5-7.3%). Symptoms loaded onto similar factors between reporters. Rates of symptom endorsement were similar between sexes, with disproportionately higher endorsement rates for youth who self-identified as sexual minority, Hispanic, Black, or Mixed race participants, and those from a disadvantaged socioeconomic background, compared to the reference ABCD sample. Youth and parent reports at 2-year showed ∼12% overlap.

**Conclusions:** ED-related concerns among historically understudied racial and sexual minority groups call for greater attention to the detection and treatment of these symptoms in these groups. Applying a transdiagnostic approach to ED symptoms can inform effective detection and intervention efforts.

**Public health statement:** Our work depicts the sociodemographic breadth of disordered eating behaviors in a large diverse sample of American youth, and underscores the importance of including multiple reporters when assessing ED symptoms in community samples of children and adolescents. Taking into consideration the diverse sociodemographic landscape of disordered eating behaviors is imperative to ensure access to care is equitable across the sexes, and racial, ethnic and socioeconomic groups.

## INTRODUCTION

Eating disorders (EDs) collectively share cardinal features, including dysregulated eating patterns and body image distortion, and often, they are characterized by a chronic, costly, and disabling illness course [1–3]. The prevalence rates of the three primary EDs, encompassing Anorexia Nervosa (AN), Bulimia Nervosa (BN), and Binge Eating Disorder (BED), range from 0.3-4% [4]. EDs disproportionately impact females as compared to males, with lifetime prevalence rates of AN reported to be ∼4% for females and 0.3% for males, and rates of BN reported to be 3% for females and 1% for males [5]. Approximately 22% of youth were recently found to endorse ED-related symptoms [6], including subthreshold ED symptoms that have been associated with significant levels of functional impairment [7]. Finally, it is not uncommon for affected individuals with EDs to exhibit diagnostic crossover over the course of their illness [3,8,9], suggesting a transdiagnostic approach may be fruitful in characterizing the landscape of ED concerns in youth.

EDs have been surrounded by misconceptions of their predominant impact on a narrowly defined group of individuals (i.e., affluent White women with low BMI) [10,11], which may pose additional barriers for patients from underrepresented minority backgrounds when seeking care when compared to those from majority groups [12]. Several reports have indicated elevated rates of disordered eating and related cognitions in sexual and gender minority individuals as compared to the general population, particularly for binge eating and body dissatisfaction [13,14]. Moreover, individuals of diverse racial and ethnic backgrounds may be at elevated risk for binge eating and other ED-related behaviors [10,12,15,16], where risk may be further compounded by social determinants of health (e.g., food insecurity, economic factors) and developmental factors [17,18]. Identifying ED symptoms in historically underrepresented individuals is imperative to determine the sociodemographic reach of EDs, which can, in turn, inform equitable early detection and intervention efforts.

Data collection from multiple informants, such as through youth self-report and their caregivers, can yield both complementary and independent information that may not be captured through the inclusion of data from a sole reporter. However, there is often low concordance between parent and youth reports of ED symptoms [19–21] and related factors such as internalizing symptoms [22]. Characterizing the degree of discordance between caretakers and their youth during early adolescence may be important in identifying clear targets for early detection efforts with family involvement, especially when considering ED-related behaviors that are less likely to be detected by caregivers.

The Adolescent Brain Cognitive Development□Study (ABCD Study^®^) provides new opportunities to probe the landscape of subthreshold ED-related concerns in youth and their caregivers in a transdiagnostic manner. First, the ABCD Study cohort includes data collection from a wide array of measures, including mental health symptoms, in a large (N=11,868 at baseline) sample of youth that reflect sociodemographic variation across the United States. Second, the study offers a valuable longitudinal study design across adolescence, capturing a neurodevelopmental window that coincides with the typical age of onset of EDs and related behaviors. The study also collects information from multiple informants, including and of relevance to this study, youth self-report and caregiver-reports. Previous studies have used the ABCD Study to estimate the prevalence of ED diagnoses in children living in the US [23–25]. However, given the age range of participants with available self-report data, they likely capture more subthreshold disordered eating concerns. Thus, the ABCD Study dataset may be better positioned to examine ED-related concerns through a transdiagnostic lens, which is supported by an increased appreciation for a dimensional view of ED symptoms and related psychiatric traits [26,27].

The aims of the current study were: 1) to identify data-driven clusters of disordered eating behaviors and related cognitions in early adolescence across parent and youth reports in a demographically-representative sample of youth in the US; 2) to characterize the sociodemographic spread of ED symptoms using identified symptom factors; and 3) to compare youth and parent-reported ED-related symptoms. We hypothesized that ED concerns would be endorsed across sexes, gender identities and sexual orientations, races, ethnicities, and socioeconomic strata. Consistent with previous work [19,21,28], we also expected parent and youth reports of ED-related concerns to diverge and for higher endorsement of symptoms from youth, particularly for more hidden behaviors, such as purging/compensatory behaviors.

## METHODS

### Participants

Data were drawn from the baseline (n=11,868) and two-year follow-up (2-year; n=10,908) timepoints included in the ABCD Study’s Curated Annual Release 5.0 (doi: 10.15154/8873-zj65; released June 2023). The ABCD Study is a longitudinal study tracking brain and behavioral development of ∼11,880 children, starting at 9-11 years of age, recruited between September 2016 and August 2018 [29,30]. The ABCD sample was recruited through epidemiologically-informed procedures to ensure demographic variation mirroring that found in the US population of 9- and 10-year-olds. Further details can be found in [29] and in Supplementary Materials. Consent (parents) and assent (children) was obtained from all participants, and the ABCD Study was approved by the appropriate institutional review boards. See **Table 1** for sample descriptives for the current study.

**Table 1.** Sample descriptives. Abbreviations: F, Female; AIAN/NHPI, American Indian American Native/Native Hawaiian and Pacific Islander; HS, High school; GED, general education development. ^1^This reflects the total sample size of ABCD included in release 5.0, but each variable within the table may have missing data. ^2^All percentages from the current row onwards are in relation to totals listed in the “N endorsed” row ^3^Baseline gender identity and sexual health measures were taken from baseline parent reports, whereas 2-year eating symptoms were matched to 3-year youth-reported GISH measures.

|  |  | ABCD Full Sample |  | Parent Report<br>Baseline |  |  | Parent Report<br>2-year |  |  | Youth Report<br>2-year |  |  |
| --- | --- | --- | --- | --- | --- | --- | --- | --- | --- | --- | --- | --- |
|  |  | Baseline | 2-year | Weight<br>Control | Weight<br>Distress | Binge | Weight<br>Control | Weight<br>Distress | Binge | Weight<br>Control | Weight<br>Distress | Binge |
| N endorsed (% full sample) |  | 11,868<sup>1</sup> | 10,908<sup>1</sup> | (3.01) | (1.52) | 596 (5.02) | 458 (4.20) | 209 (1.92) | 673 (6.17) | (7.13) | 243 (2.23) | 794 (7.28) |
| Years (Range) |  | 11.08) | (10.58-14) | (8.92-11) | 11) | 11) | 13.67) | 13.75) | 13.83) | (10.67- | 13.67) | (10.75- |
| Sex, gender<br>identity and<br>sexual<br>orientation<sup>2</sup> | Sex at birth (F, %) | 5677<br>(47.83) | 5181<br>(47.5) | 171<br>(47.9) | 96<br>(53.33) | 254<br>(42.62) | 220 (48.03) | 105 (50.48) | 299 (44.56) | 357<br>(46.12) | 158 (65.56) | 382 (48.11) |
|  | Gender Minority<br>(Yes/Maybe, %<sup>3</sup>) | 135 (1.14) | 344 (3.43) | 2 (0.56) | 2 (1.12) | 6 (1.01) | 5 (1.09) | 2 (0.96) | 6 (0.89) | 25 (3.58) | 21 (9.42) | 40 (5.56) |
|  | Sexual Minority<br>(Yes/Maybe, %<sup>3</sup>) | 904 (7.63) | 804 (7.99) | 27 (7.58) | 17 (9.5) | 58 (9.73) | 26 (5.69) | 15 (7.25) | 65 (9.69) | 65 (9.27) | 52 (23.21) | 125 (17.24) |
| Race (%) | AIAN/NHPI | 78 (0.67) | 66 (0.61) | 2 (0.58) | 0 (0) | 3 (0.51) | 4 (0.88) | 1 (0.48) | 5 (0.76) | 11 (1.45) | 3 (1.29) | 7 (0.9) |
|  | Asian | 275 (2.35) | 250 (2.32) | 8 (2.33) | 3 (1.69) | 11 (1.88) | 12 (2.65) | 3 (1.45) | 17 (2.57) | 7 (0.92) | 2 (0.86) | 7 (0.9) |
|  | Black | (15.98) | (15.09) | (27.62) | (21.91) | (20.82) | 86 (19.03) | 24 (11.59) | 88 (13.29) | (20.66) | 61 (26.29) | 138 (17.83) |
|  | Mixed | (12.26) | (12.27) | (16.57) | (18.54) | 86 (14.68) | 68 (15.04) | 33 (15.94) | 116 (17.52) | (11.97) | 27 (11.64) | 94 (12.14) |
|  | Other | 525 (4.49) | 464 (4.31) | 21 (6.1) | 14 (7.87) | 35 (5.97) | 27 (5.97) | 11 (5.31) | 36 (5.44) | 55 (7.24) | 10 (4.31) | 35 (4.52) |
|  | White | (64.26) | (65.39) | (46.8) | 89 (50) | (56.14) | 255 (56.42) | 135 (65.22) | 400 (60.42) | (57.76) | 129 (55.6) | 493 (63.7) |
| Ethnicity | Hispanic (Yes, %) | (20.57) | (19.93) | 112 (32) | (33.71) | (30.41) | 129 (28.73) | 45 (22.06) | 177 (26.82) | (28.33) | 60 (25) | 194 (24.84) |
| Parental highest<br>education (%) | <HS Diploma | 593 (5) | 488 (4.48) | (10.67) | 16 (8.94) | 38 (6.4) | 37 (8.11) | 14 (6.76) | 40 (5.98) | 45 (5.84) | 19 (7.88) | 42 (5.3) |
|  | HS Diploma/GED | 1132 (9.55) | (9.27) | (14.33) | (11.17) | 85 (14.31) | 47 (10.31) | 14 (6.76) | 75 (11.21) | (14.14) | 37 (15.35) | 81 (10.21) |
|  | Some College | (25.93) | (24.3) | (33.71) | (36.31) | (34.34) | 146 (32.02) | 66 (31.88) | 177 (26.46) | (31.91) | 77 (31.95) | 234 (29.51) |
|  | Bachelor | (25.42) | (25.83) | (22.47) | (21.79) | (20.88) | 96 (21.05) | 49 (23.67) | 151 (22.57) | (21.79) | 49 (20.33) | 224 (28.25) |
|  | Postgrad degree | 4042 (34.1) | (36.12) | (18.82) | (21.79) | (24.07) | 130 (28.51) | 64 (30.92) | 226 (33.78) | (26.33) | 59 (24.48) | 212 (26.73) |
| Income (%) | <50K | (29.69) | (24.85) | (49.85) | 68 (42.5) | (44.16) | 157 (38.39) | 64 (33.16) | 201 (32.68) | (33.67) | 82 (38.14) | 211 (29.02) |
|  | ≥50K & <100K | (28.27) | (26.86) | 83 (25.7) | (28.75) | (27.46) | 110 (26.89) | 50 (25.91) | 166 (26.99) | (29.94) | 73 (33.95) | 223 (30.67) |
|  | ≥100K | (42.03) | (48.3) | (24.46) | (28.75) | (28.39) | 142 (34.72) | 79 (40.93) | 248 (40.33) | (36.39) | 60 (27.91) | 293 (40.3) |

#### ED symptoms and factors

Current ED symptoms related to disordered eating behaviors and cognitions were drawn from the ED Module of the KSADS, a semi-structured, self-administered, computerized version of the Kiddie Schedule for Affective Disorders and Schizophrenia for School-Aged Children (KSADS-5) [31,32]. Additional details on the KSADS for use in ABCD can be found in [33] and in Supplementary Material. Data were collected from both parent/caregivers reporting on their youth (hereafter referred to as parent-report), and youth self-reports. Parent-report data are available for both baseline and 2-year timepoints, whereas the youth completed the ED Module from 2-year follow-up onwards.

#### Sociodemographic variables

Sociodemographic variables included age, sex at birth, race, ethnicity, parental highest education and household income, as described in [34], with additional details in Supplementary Materials. Additionally, we incorporated gender identity and sexual health data [35] from KSADS background items on Gender Identity and Sexual Orientation [36] to capture youth that identified as a sexual and/or gender minority. For sexual minorities, we adapted the definitions described by Nagata and colleagues [37]. See supplementary material for detailed information.

#### Data Analyses

All statistical analyses were carried out with *R* version 4.1.3.

##### Factor analysis

To derive symptom factors, an exploratory tetrachoric factor analysis using an oblique rotation was applied to eight cognitive and behavioral symptom data listed in Figures 1 and 2, for parent baseline, parent 2-year, and youth 2-year reports. Given the familial structure of the ABCD Study, one participant was randomly selected if any siblings/twins were present, such that the factor analysis was carried out on singletons only. The factor analysis was carried out on data where at least one symptom was endorsed through i) parent-reported baseline data (*n*=792); ii) parent-reported two-year data (*n*=939); and iii) youth-reported two-year data (*n*=1233). The optimal factor solution was determined through assessing the proportion of variance explained and visualization of scree plots (Supplementary Figure 1), comparing model fits with the Bayesian Information Criterion (BIC) score, and selecting a factor that minimized the root mean square residual (RMSR).

**Figure 1.**
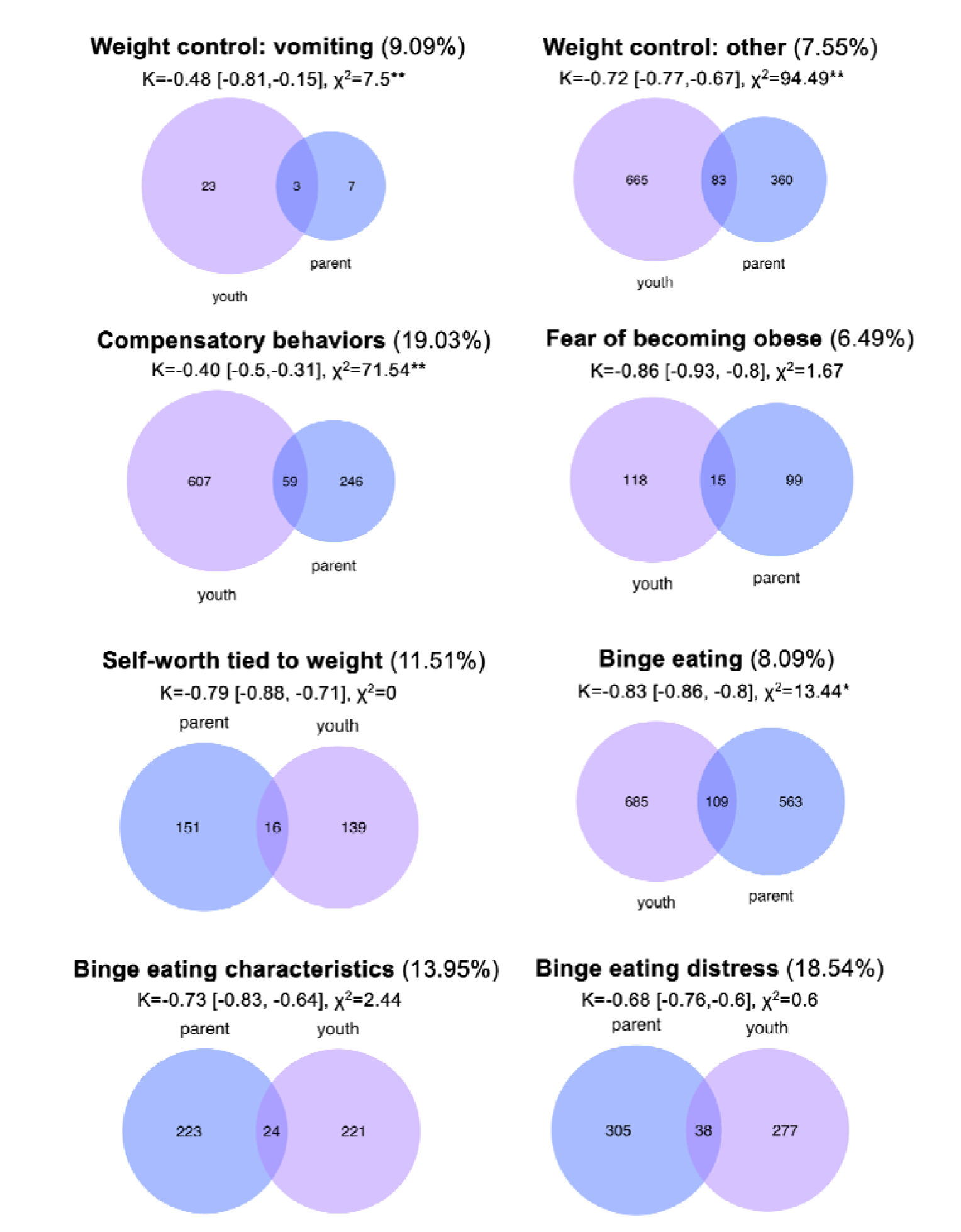
Venn Diagrams comparing the number of participants endorsing the listed symptom at 2-year between youth (purple) and parent-reports (blue). Percent overlap between youth and parents is included in each of the item headers in parentheses, as well as Cohen’s kappa (with 95% confidence intervals) and McNemar’s chi-squared test statistics, to test for statistical overlap and significant differences between reporters, respectively. McNemar’s exact test was used for the vomiting item, given the smaller sample size. On average, parents and youth showed 11.78% overlap across these items. Only cases where the question was asked to both reporters or at both timepoints were retained. Note, parents and youth were included in these diagrams only if both reporters were asked the question. Asterisks refer to significance level of p-values for McNemar’s test statistic: **Bonferroni-corrected p<0.00625; *Nominally significant at p<0.05.

**Figure 2.**
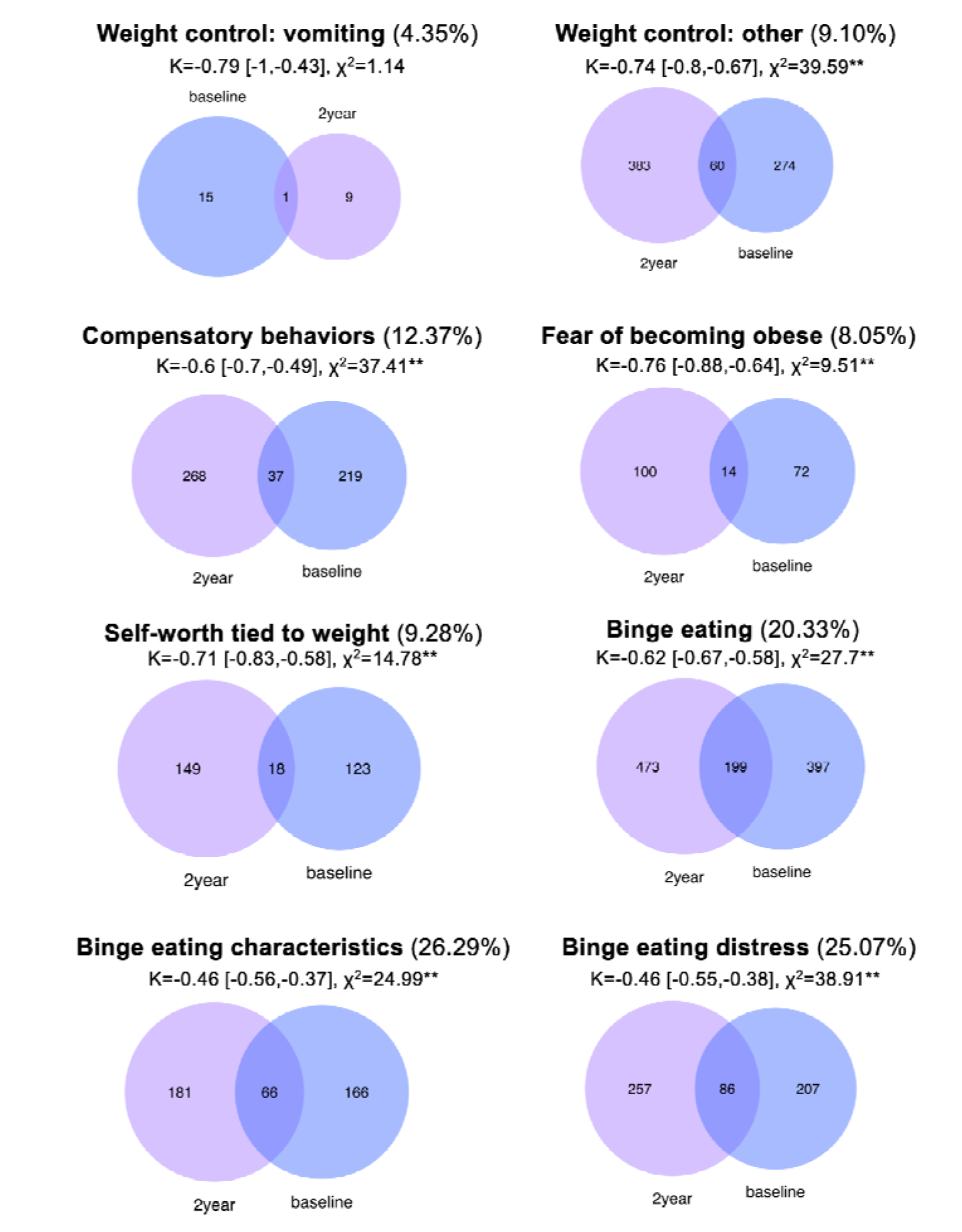
Venn Diagrams comparing the number of parents reporting on their youth endorsing the listed symptom at baseline (blue) and 2-year (purple). Percent overlap between timepoints is included in each of the item headers in parentheses, as well as Cohen’s kappa (with 95% confidence intervals) and McNemar’s test statistics, to test for statistical overlap and significant differences between reporters, respectively. McNemar’s exact test was used for the vomiting item, given the smaller sample size. On average, parents showed 14.36% overlap across items between the two timepoints. Note, baseline and two-year data were included in these diagrams only if parents were asked the question at the both timepoints. Asterisks refer to significance level of p-values for McNemar’s test statistic:**Bonferroni-corrected p<0.00625.

##### Analyses with sociodemographic variables

Log odds ratios (LORs) with corresponding Wald’s statistics and confidence intervals were calculated for each sociodemographic categorical variable (i.e., sex, gender/sexual minority status, race, ethnicity, and socioeconomic resources) to compare the probability of being in a particular category given symptom endorsement within an ED factor to the probability of being in that same category in the entire ABCD sample.

##### Comparing symptoms between informants and across time

Cohen’s kappa and McNemar’s tests were used to test for either significant overlap (Cohen’s kappa) or significant differences (McNemar’s test) between parent and youth two-year reports, as well as between parent baseline and parent two-year reports. Percent overlap between either reporter or timepoint were also calculated. Tests were carried out only on participants that had a symptom endorsed by either parent or youth (or at minimum one timepoint) to ensure that overlap metrics would not be inflated by agreement of the absence of behavior, which characterizes most of the ABCD sample. Further, only cases where the question was asked to both reporters or at both timepoints were retained.

## RESULTS

### Factor analysis results

The factor analysis generally yielded a stronger 4-factor solution, where the vomiting item (*ksads_13_71*) loaded onto its own factor. However, given the small sample size of participants endorsing this item, a 3-factor solution was used for all analyses in the manuscript, reflecting “weight distress”, “weight control”, and “binge eating” (Supplementary Figures 1 and 2). Items loaded similarly on each factor across timepoints and reporters, with the exception of the vomiting item loading more onto the weight distress factor for youth-2 year reports, and relatively equally on weight distress and weight control for parent 2-year reports. The three-factor solution and additional statistics are displayed in Supplementary Figure 1.

### Sociodemographic spread of current ED symptoms and factors

Sociodemographic descriptives per informant and timepoint are included in **Table 1**, organized by the three resultant factors, as described above. As a comparison point, the sociodemographic spread of the 11,868 and 10,908 youth with baseline and two-year data, respectively, included in release 5.0 is included in **Table 1**. Across timepoints and reporters, factor symptom endorsement ranged from 1.52% to 7.28%, with the lowest endorsement for the weight distress factor, and highest for youth-reported binge eating. LORs comparing odds of being in a particular sociodemographic category given endorsement of at least one of the ED factor indicators compared to the reference ABCD sample, 95% confidence intervals, and corresponding Wald’s statistics are listed in **Tables 2-4**. A summary of key significant results are detailed below.

**Table 2.**
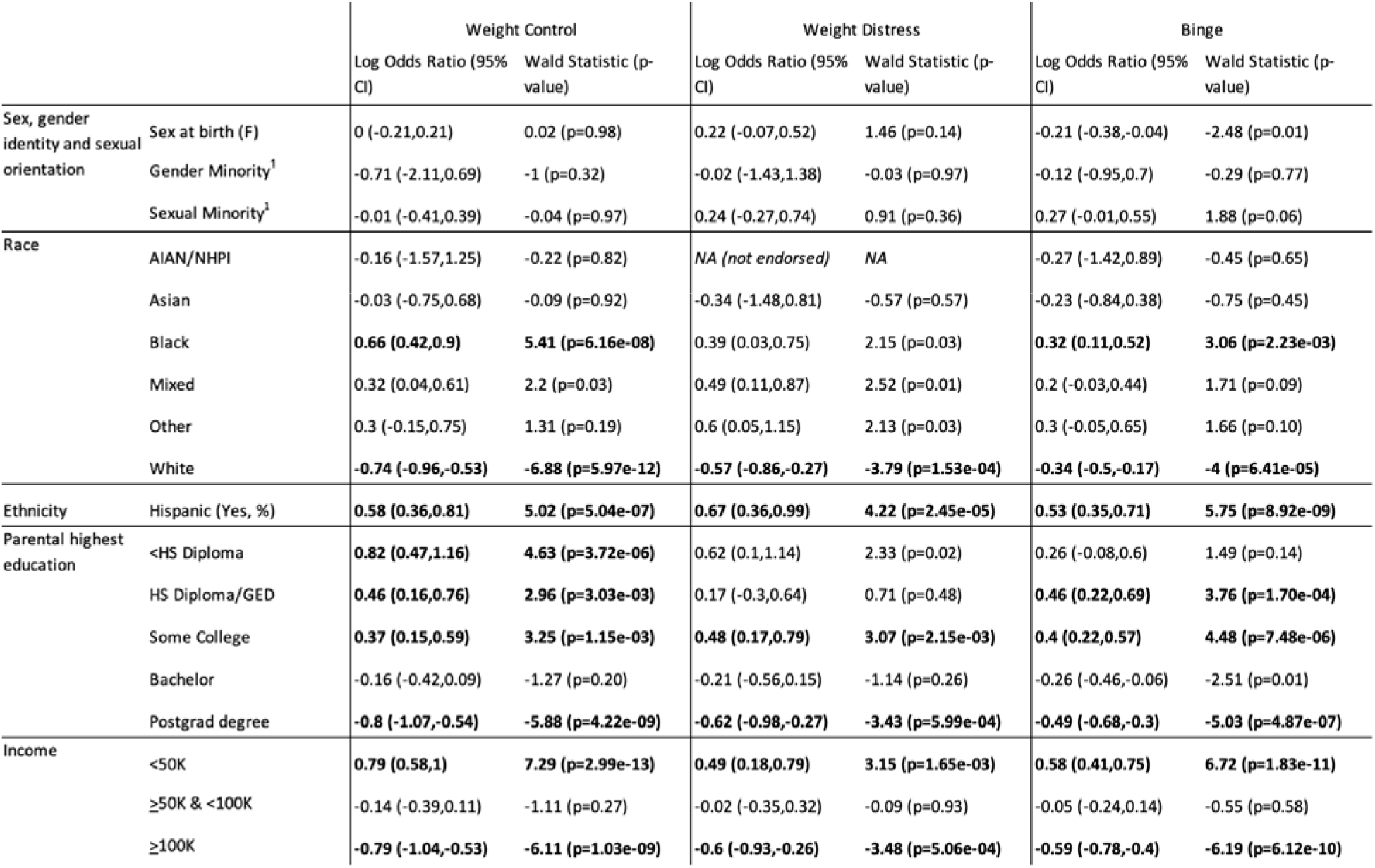
Baseline Parent Report: Log Odds Ratios & corresponding statistics comparing ED groups to full ABCD sample. Positive log odds ratios reflect that participants endorsing a given ED symptom have higher odds of being in a given sociodemographic group compared to the reference ABCD population, whereas negative ratios indicate that participants with ED symptoms are less likely to be in a given sociodemographic category. Bolded cells reflect significantly different proportions at p<0.01. Abbreviations: F, Female; AIAN/NHPI, American Indian American Native/Native Hawaiian and Pacific Islander; HS, High school; GED, general education development. ^1^Baseline gender identity and sexual health measures were inferred from baseline parent reports.

**Table 3.** Two-year Parent Report: Log Odds Ratios & corresponding statistics comparing ED groups to full ABCD sample. Positive log odds ratios reflect that participants endorsing a given ED symptom have higher odds of being in a given sociodemographic group compared to the reference ABCD population, whereas negative ratios indicate that participants with ED symptoms are less likely to be in a given sociodemographic category. Bolded cells reflect significantly different proportions at p<0.01. Abbreviations: F, Female; AIAN/NHPI, American Indian American Native/Native Hawaiian and Pacific Islander; HS, High school; GED, general education development. ^1^2-year eating symptoms were matched to 3-year youth-reported gender identity and sexual health measures.

|  |  | Weight Control |  | Weight Distress |  | Binge |  |
| --- | --- | --- | --- | --- | --- | --- | --- |
|  |  | Log Odds Ratio (95% CI) | Wald Statistic (p-value) | Log Odds Ratio (95% CI) | Wald Statistic (p-value) | Log Odds Ratio (95% CI) | Wald Statistic (p-value) |
| Sex, gender identity and sexual orientation | Sex at birth (F) | 0.02 (-0.17,0.21) | 0.23 (p=0.82) | 0.11 (-0.16,0.38) | 0.79 (p=0.43) | -0.12 (-0.28,0.03) | -1.55 (p=0.12) |
|  | Gender Minority<sup>1</sup> | -1.08 (-1.97,-0.19) | -2.39 (p=0.02) | -1.22 (-2.61,0.18) | -1.7 (p=0.09) | **-1.29 (-2.1,-0.48)** | **-3.11 (p=1.87e-03)** |
|  | Sexual Minority<sup>1</sup> | -0.28 (-0.68,0.12) | -1.36 (p=0.17) | -0.03 (-0.56,0.5) | -0.11 (p=0.92) | 0.3 (0.03,0.56) | 2.18 (p=0.03) |
| Race | AIAN/NHPI | 0.37 (-0.64,1.38) | 0.71 (p=0.47) | -0.24 (-2.22,1.74) | -0.23 (p=0.82) | 0.21 (-0.71,1.12) | 0.44 (p=0.66) |
|  | Asian | 0.14 (-0.45,0.72) | 0.46 (p=0.65) | -0.48 (-1.62,0.67) | -0.81 (p=0.42) | 0.1 (-0.4,0.6) | 0.39 (p=0.69) |
|  | Black | 0.28 (0.04,0.52) | 2.28 (p=0.02) | -0.3 (-0.73,0.13) | -1.36 (p=0.17) | -0.15 (-0.38,0.08) | -1.28 (p=0.20) |
|  | Mixed | 0.24 (-0.03,0.5) | 1.75 (p=0.08) | 0.31 (-0.07,0.69) | 1.61 (p=0.11) | **0.41 (0.21,0.62)** | **3.9 (p=9.72e-05)** |
|  | Other | 0.34 (-0.06,0.74) | 1.68 (p=0.09) | 0.22 (-0.39,0.84) | 0.71 (p=0.48) | 0.24 (-0.11,0.59) | 1.35 (p=0.18) |
|  | White | **-0.37 (-0.56,-0.18)** | **-3.83 (p=1.28e-04)** | 0 (-0.28,0.29) | 0.03 (p=0.97) | **-0.21 (-0.37,-0.06)** | **-2.65 (p=8.11e-03)** |
| Ethnicity | Hispanic (Yes, %) | **0.47 (0.26,0.68)** | **4.4 (p=1.07e-05)** | 0.11 (-0.22,0.45) | 0.66 (p=0.51) | **0.38 (0.2,0.55)** | **4.13 (p=3.59e-05)** |
| Parental highest education | <HS Diploma | **0.63 (0.28,0.98)** | **3.54 (p=3.94e-04)** | 0.43 (-0.12,0.98) | 1.52 (p=0.13) | 0.3 (-0.03,0.63) | 1.77 (p=0.08) |
|  | HS Diploma/GED | 0.11 (-0.19,0.42) | 0.72 (p=0.47) | -0.35 (-0.9,0.19) | -1.26 (p=0.21) | 0.21 (-0.04,0.45) | 1.63 (p=0.10) |
|  | Some College | **0.38 (0.18,0.58)** | **3.69 (p=2.23e-04)** | 0.37 (0.07,0.66) | 2.43 (p=0.02) | 0.11 (-0.07,0.29) | 1.2 (p=0.23) |
|  | Bachelor | -0.27 (-0.5,-0.04) | -2.31 (p=0.02) | -0.13 (-0.45,0.2) | -0.77 (p=0.44) | -0.18 (-0.37,0) | -1.93 (p=0.05) |
|  | Postgrad degree | **-0.35 (-0.56,-0.15)** | **-3.34 (p=8.25e-04)** | -0.25 (-0.54,0.05) | -1.62 (p=0.11) | -0.11 (-0.27,0.06) | -1.3 (p=0.19) |
| Income | <50K | **0.57 (0.37,0.77)** | **5.63 (p=1.82e-08)** | **0.4 (0.1,0.7)** | **2.65 (p=8.12e-03)** | **0.37 (0.19,0.54)** | **4.19 (p=2.74e-05)** |
|  | ≥50K & <100K | -0.03 (-0.25,0.18) | -0.3 (p=0.76) | -0.04 (-0.36,0.28) | -0.24 (p=0.81) | 0 (-0.18,0.18) | 0.01 (p=0.99) |
|  | ≥100K | **-0.57 (-0.77,-0.37)** | **-5.56 (p=2.74e-08)** | -0.27 (-0.55,0.01) | -1.87 (p=0.06) | **-0.31 (-0.47,-0.15)** | **-3.78 (p=1.60e-04)** |

**Table 4.** Two-year Youth Report: Log Odds Ratios & corresponding statistics comparing ED groups to full ABCD sample. Positive log odds ratios reflect that participants endorsing a given ED symptom have higher odds of being in a given sociodemographic group compared to the reference ABCD population, whereas negative ratios indicate that participants with ED symptoms are less likely to be in a given sociodemographic category. Bolded cells reflect significantly different proportions at p<0.01. Abbreviations: F, Female; AIAN/NHPI, American Indian American Native/Native Hawaiian and Pacific Islander; HS, High school; GED, general education development. ^1^2-year eating symptoms were matched to 3-year youth-reported gender identity and sexual health measures.

|  |  | Weight Control |  | Weight Distress |  | Binge |  |
| --- | --- | --- | --- | --- | --- | --- | --- |
|  |  | Log Odds Ratio (95% CI) | Wald Statistic (p-value) | Log Odds Ratio (95% CI) | Wald Statistic (p-value) | Log Odds Ratio (95% CI) | Wald Statistic (p-value) |
| Sex, gender identity and sexual orientation | Sex at birth (F) | -0.06 (-0.21,0.08) | -0.87 (p=0.38) | **0.72 (0.45,0.99)** | **5.3 (p=1.16e-07)** | 0.02 (-0.12,0.17) | 0.33 (p=0.74) |
|  | Gender Minority<sup>1</sup> | 0.02 (-0.39,0.43) | 0.09 (p=0.93) | **1.07 (0.61,1.53)** | **4.54 (p=5.57e-06)** | **0.49 (0.15,0.82)** | **2.85 (p=4.37e-03)** |
|  | Sexual Minority<sup>1</sup> | 0.14 (-0.13,0.4) | 1.01 (p=0.31) | **1.23 (0.92,1.54)** | **7.66 (p=1.91e-14)** | **0.85 (0.65,1.06)** | **8.2 (p=2.22e-16)** |
| Race | AIAN/NHPI | **0.86 (0.21,1.5)** | **2.61 (p=8.94e-03)** | 0.72 (-0.44,1.88) | 1.21 (p=0.23) | 0.38 (-0.4,1.16) | 0.95 (p=0.34) |
|  | Asian | -0.95 (-1.7,-0.19) | -2.47 (p=0.01) | -1.04 (-2.44,0.36) | -1.46 (p=0.14) | -0.97 (-1.72,-0.22) | -2.52 (p=0.01) |
|  | Black | **0.37 (0.19,0.55)** | **3.96 (p=7.63e-05)** | **0.65 (0.36,0.95)** | **4.33 (p=1.50e-05)** | 0.19 (-0.01,0.38) | 1.9 (p=0.06) |
|  | Mixed | -0.04 (-0.26,0.19) | -0.33 (p=0.74) | -0.1 (-0.5,0.31) | -0.47 (p=0.64) | -0.02 (-0.25,0.2) | -0.22 (p=0.83) |
|  | Other | **0.54 (0.25,0.83)** | **3.64 (p=2.71e-04)** | -0.03 (-0.67,0.61) | -0.11 (p=0.92) | 0.04 (-0.31,0.39) | 0.21 (p=0.84) |
|  | White | **-0.34 (-0.49,-0.19)** | **-4.51 (p=6.64e-06)** | **-0.47 (-0.73,-0.22)** | **-3.63 (p=2.78e-04)** | -0.1 (-0.25,0.05) | -1.36 (p=0.17) |
| Ethnicity | Hispanic (Yes, %) | **0.46 (0.29,0.62)** | **5.46 (p=4.77e-08)** | 0.29 (0,0.59) | 1.93 (p=0.05) | **0.28 (0.11,0.45)** | **3.22 (p=1.30e-03)** |
| Parental highest education | <HS Diploma | 0.27 (-0.04,0.59) | 1.69 (p=0.09) | 0.59 (0.12,1.07) | 2.44 (p=0.01) | 0.18 (-0.15,0.5) | 1.07 (p=0.29) |
|  | HS Diploma/GED | **0.47 (0.26,0.68)** | **4.32 (p=1.60e-05)** | **0.57 (0.21,0.92)** | **3.11 (p=1.85e-03)** | 0.11 (-0.13,0.35) | 0.88 (p=0.38) |
|  | Some College | **0.37 (0.21,0.52)** | **4.58 (p=4.75e-06)** | **0.37 (0.1,0.64)** | **2.65 (p=8.00e-03)** | **0.27 (0.11,0.42)** | **3.28 (p=1.02e-03)** |
|  | Bachelor | **-0.23 (-0.41,-0.06)** | **-2.59 (p=9.65e-03)** | -0.32 (-0.64,0) | -1.98 (p=0.05) | 0.12 (-0.04,0.28) | 1.5 (p=0.13) |
|  | Postgrad degree | **-0.47 (-0.63,-0.3)** | **-5.58 (p=2.46e-08)** | **-0.56 (-0.86,-0.27)** | **-3.74 (p=1.82e-04)** | **-0.44 (-0.6,-0.28)** | **-5.29 (p=1.22e-07)** |
| Income | <50K | **0.38 (0.22,0.54)** | **4.7 (p=2.64e-06)** | **0.54 (0.28,0.81)** | **3.96 (p=7.46e-05)** | 0.2 (0.04,0.37) | 2.43 (p=0.01) |
|  | ≥50K & <100K | 0.12 (-0.05,0.28) | 1.39 (p=0.17) | 0.27 (-0.01,0.55) | 1.92 (p=0.05) | 0.18 (0.02,0.34) | 2.16 (p=0.03) |
|  | ≥100K | **-0.5 (-0.65,-0.34)** | **-6.29 (p=3.21e-10)** | **-0.89 (-1.18,-0.59)** | **-5.91 (p=3.39e-09)** | **-0.31 (-0.46,-0.16)** | **-4.05 (p=5.07e-05)** |

#### Sex, gender identity, and sexual orientation

Across both reporters and timepoints, symptoms were endorsed by both males and females, with more female-at-birth youth endorsing higher weight distress at 2-year (65.6%) compared to the full ABCD sample proportion of females (47.5%). The most striking differences in gender and sexual minority youth proportions endorsing ED symptoms compared to the full sample emerged for 2-year youth reports, where i) disproportionately more gender minority youth endorsed weight distress (9.42%) and binge symptoms (5.56%) compared to the 3.43% of gender minority youth in the full 2-year sample; and ii) disproportionately more sexual minority youth endorsed weight distress (23.21%) and binge symptoms (17.24%), compared to the 7.99% of sexual minority youth in the full sample. With the exception of 2-year parent-reported gender minority status and binge eating (0.89%), parent reports did not reveal significant differences in proportions of gender (baseline, 2-year) or sexual minority (baseline) youth endorsing ED symptoms compared to the full ABCD sample.

#### Race and ethnicity

Across reporters and timepoints, a pattern emerged of disproportionately higher rates of all three symptom factors among participants who self-identified as Black, Other/Mixed race, and Hispanic compared to the full ABCD sample (15-16%, 17%, 20-21% respectively). The largest differences emerged in higher proportions of i) Black youth with weight control symptoms (baseline parent report: 27.62%; youth report: 20.66%), ii) Mixed race youth with 2-year parent reported binge symptoms (23.72%); and iii) Hispanic-identifying youth with baseline parent-reported binge symptoms (33.71%) and with 2-year youth reported weight control symptoms (28.33%).

#### Socioeconomic resources

Across reporters and timepoints, an overall trend emerged of disproportionately higher rates of ED symptoms in youth coming from households with lower levels of parental education and lower income (comprising ∼15% and 25-30%, respectively, in the full ABCD sample). Notable differences included higher proportions of i) parent-reported weight control symptoms in youth having a parent without a high school diploma (baseline: 10.67%; 2-year: 8.11%), compared to ∼5% of participants with a similar parental education level in the full ABCD sample; ii) baseline parent-reported binge symptoms (14.31%), and 2-year youth-reported weight control (14.14%) and weight distress (15.35%) symptoms were higher in youth with parents with a high school diploma/GED, compared to ∼9% of participants in this parental education category in the full ABCD sample; and iii) for participants coming from a household income of <$50,000 annually, disproportionately higher weight distress, weight control and binge symptoms (32.7-49.9%) for both baseline and 2-year parent reports, as well as disproportionately higher youth-reported weight control (33.67%) and weight distress (38.14%) symptoms, compared to ∼30 and 25% of baseline and 2-year ABCD samples, respectively, in this household income category.

### Comparing symptoms between informants and across time

The proportion of overlap between youth and parent report at 2-year was on average 11.78% across individual items, ranging from 6.49% overlap for “fear of becoming obese”, and 19.03% for compensatory behaviors (**Figure 1**). Higher rates of symptom endorsement were found for 2-year data compared to baseline for parent reports. Youth reported higher rates of individual symptom endorsement compared to parent 2-year reports. Proportion of overlap between baseline and 2-year parent reports was on average 14.36%, ranging from 4.35% for vomiting and 26.29% for characteristics binge eating (**Figure 2**). For the symptom factors, parents generally reported higher ED symptoms for their youth from baseline to 2-year across all three factors. Cohen’s kappa and McNemar’s chi-squared statistics are reported in Figures 1 and 2. Significant and negative Cohen’s kappa values indicate a low level of agreement between parent and youth reports, and parent baseline and parent two-year reports. This result is also confirmed by McNemar’s tests.

## DISCUSSION

Our results depict the sociodemographic breadth of disordered eating behaviors and cognitions in a large diverse sample of American youth, and underscore the importance of including multiple reporters when assessing ED symptoms in community samples of children and adolescents. Across both parent and youth reports, ED concerns are relatively equally distributed across sexes, with disproportionately higher endorsement in sexual and/or gender minority groups, Black, Mixed race and Hispanic-identifying youth, and youth with lower socioeconomic resources. Parent-reported ED symptoms generally increased from baseline to two-year [38], with low concordance on symptom endorsement between youth and parents/caregivers in early adolescence, consistent with previous reports [19,21]. These results suggest that many of these behaviors may be hidden or undetected by family members of affected youth, which holds important implications for early detection and intervention efforts at a critical time period in the development of youth.

The ABCD Study has previously been leveraged to map the sociodemographic spread of binge eating disorders and binge-related behaviors using parent report and baseline sexual identity data [14]. We complement and extend these results to youth reports and sexual identity data collected in adolescence, and importantly, we map sociodemographic and behavioral variables to symptom factors that underlie restrictive EDs as well. Many ED symptoms overlap across diagnostic categories; thus our symptom factor approach provides a transdiagnostic perspective that may aid in risk and biomarker discovery compared to traditional diagnostic approaches.

Taking into consideration the diverse sociodemographic landscape of ED-related symptoms is imperative to ensure access to care is equitable across the sexes, and racial, ethnic and socioeconomic groups. It has been found that healthcare resources are underutilized by men and racial/ethnic minorities [12], as well as sexual minority youth [39]. Intersectionality may amplify such inequities; for instance, disproportionately high levels of ED symptoms have been found in individuals identifying with both a sexual and racial minority group [18]. A recent investigation using ABCD Study data also found that weight-based discrimination and disordered eating were higher in youth who experienced racial/ethnic and sexual orientation discrimination [40]. This underscores the importance of diversifying efforts in ED research to move past convenience samples and historically overrepresented groups (i.e. White women from affluent backgrounds) and understand the complex interplay between various social determinants of health in the development of ED-related concerns.

In the current investigation, there was a high degree of discordance between parent and youth reports at the late childhood/early adolescent time period for which data from both reporters were available. Previous work in this area has suggested that some symptoms may be endorsed at higher rates by youth (e.g., binge eating and/or purging) [19,21,28], but there are other constructs where youth may lack insight (e.g., denial of being ill, particularly in restrictive EDs such as AN) and parent/caregiver reports may be more elucidating [21]. Concordance between parent and youth reports may also differ by age of the affected youth [41,42]. The discordance we report emphasizes the importance of better education and detection of ED-related concerns for both parents and youth earlier in life, so that these symptoms can be more clearly identified. Future investigations would benefit from merging information across informants, and potentially incorporate objective measures of physical health that may complement and lend increased confidence of the symptom endorsement data collected.

The ABCD Study provides a rich data resource to explore risk factors for psychopathology, but readers should keep in mind that psychiatric symptoms in this study are not assessed by a clinician. Inclusion of more longitudinal data from ABCD will be imperative, especially as more youth meet diagnostic criteria for an ED. Youth self-reports on ED symptoms were not obtained at baseline, as some of these questions were not considered to be developmentally appropriate for youth to answer, alongside concerns around variability in language skills and retrospective reporting [33]. Finally, as ABCD represents a community based sample, future work should examine if differences in symptom endorsement across informants generalize to clinically-ascertained groups and across assessments.

Our work leverages the longitudinal and prospective study design of the ABCD Study to map the landscape of ED symptoms in early adolescence across reporters and timepoints in a transdiagnostic framework. We shed a spotlight on the prevalence of ED symptoms in historically understudied minority and under-resourced groups, and the discordance in symptoms between reporters, which can help motivate future efforts to educate caretakers and youth on the manifestation of EDs and identify better markers for early ED detection.

## Data Availability

All data produced in the present work are contained in the manuscript or are available upon reasonable request to the authors.

http://dx.doi.org/10.15154/8873-zj65

## ACKNOWLEDGMENTS/FUNDING

The authors would like to thank the research participants and staff involved in data collection of the Adolescent Brain Cognitive Development (ABCD) Study data. The ABCD Study is a multisite, longitudinal study designed to recruit more than 10,000 children ages 9 and 10 and follow them over 10 years into early adulthood. The ABCD Study is supported by the National Institutes of Health (NIH) and additional federal partners under award numbers U01DA041048, U01DA050989, U01DA051016, U01DA041022, U01DA051018, U01DA051037, U01DA050987, U01DA041174, U01DA041106, U01DA041117, U01DA041028, U01DA041134, U01DA050988, U01DA051039, U01DA041156, U01DA041025, U01DA041120, U01DA051038, U01DA041148, U01DA041093, U01DA041089, U24DA041123, and U24DA041147. A full list of supporters is available at https://abcdstudy.org/federal-partners.html. A listing of participating sites and a complete listing of the study investigators can be found at https://abcdstudy.org/study-sites/. ABCD consortium investigators designed and implemented the study and/or provided data but did not necessarily all participate in analysis or writing of this report. This manuscript reflects the views of the authors and may not reflect the opinions or views of the NIH or ABCD consortium investigators. The ABCD data repository grows and changes over time. Data were drawn from the NIMH Data Archive ABCD Collection Release 5.0 (DOI: 10.15154/8873-zj65).

This work was supported by the National Institutes of Mental Health, award number 1K99MH132886 (CM). MLW is supported by a Wellcome Trust Sir Henry Wellcome Postdoctoral Fellowship (224107/Z/21/Z).

## COMPETING INTERESTS

The authors have no competing interests to disclose.

## Notes

### Competing Interest Statement

The authors have declared no competing interest.

### Funding Statement

This work was supported by the National Institutes of Mental Health, K99MH132886, awarded to CM.

### Author Declarations

The study used openly available human data, originally located within the NIMH Data Archive ABCD Collection Release 5.0 (DOI: 10.15154/8873-zj65).

### Summary of Updates

Manuscript now focuses on sociodemographic correlates of eating disorder related symptoms based on parent and youth reports. The premorbid factor analysis included in originally uploaded preprint will be included in a new manuscript.

