## Supplementary for "Sociodemographic correlates of parent and youth-reported eating disorder symptoms in the Adolescent Brain Cognitive Development Study"

**SUPPLEMENTARY METHODS**

*Participants*.

Data were drawn from the baseline (n=11,868) and two-year follow-up (2-year; n=10,908) timepoints included in the ABCD Study’s Curated Annual Release 5.0 (doi: 10.15154/8873-zj65; released June 2023). The ABCD Study is a longitudinal study tracking brain and behavioral development of ~11,880 children, starting at 9-11 years of age, recruited between September 2016 and August 2018 [1,2]. The ABCD sample was recruited through epidemiologically-informed procedures to ensure demographic variation mirroring that found in the US population of 9- and 10-year-olds. As described in [1], a probability sampling of schools was conducted within the defined catchment areas of the study’s nationally distributed set of 21 recruitment sites. All children in each sampled school were invited to participate following classroom-based presentations, distribution of study materials and telephone screening for eligibility. Exclusions included common MRI contraindications (e.g., cardiac pacemakers and defibrillators, internal pacing wires, cochlear and metallic implants and Swan–Ganz catheters), inability to understand or speak English fluently, uncorrected vision, hearing or sensorimotor impairments, a history of major neurological disorders, gestational age <28 weeks, birth weight <1,200 g, birth complications that resulted in hospitalization for more than 1 month, current diagnosis of schizophrenia, moderate or severe autism spectrum disorder, a history of traumatic brain injury or unwillingness to complete assessments. The ABCD sample also includes 2,105 monozygotic and dizygotic twins. Consent (parents) and assent (children) was obtained from all participants, and the ABCD Study was approved by the appropriate institutional review boards. See **Table 1** of the main manuscript for sample descriptives for the current study.

*ED symptoms and factors.*

Current ED symptoms related to disordered eating behaviors and cognitions were drawn from the ED Module of the KSADS, a semi-structured, self-administered, computerized version of the Kiddie Schedule for Affective Disorders and Schizophrenia for School-Aged Children (KSADS-5) [3,4]. There is high percent agreement (between 88-86% for various diagnostic categories) between diagnoses derived from this self-administered computerized version and clinician-administered pen-and-paper KSADS-5, as well as kappa reliability estimated within the good to excellent range [3]). A Spanish version was also administered as needed, which has been shown to have high inter-rater reliability (κ>0.7) [5]. Research Assistants had extensive training to support participants as they completed this assessment, including additional guidance from the developer of the computerized version, Dr. Joan Kaufman [3], and ongoing WEBEX trainings with key senior investigators in the ABCD Study consortium with extensive background in adolescent psychiatry. Additional details on administration, validity and reliability of the KSADS for use in ABCD can be found in [6]. Data were collected from both parent/caregivers reporting on their youth (hereafter referred to as parent-report), and youth self-reports. Parent-report data are available for both baseline and 2-year timepoints, whereas the youth completed the ED Module from 2-year follow-up onwards.

*Sociodemographic variables*.

Sociodemographic variables included age, sex at birth, race, ethnicity, parental highest education and household income, as described in [7]. Reported race had 22 options, which were collapsed into six groups for descriptive purposes: American Indian American Native/Native Hawaiian and Pacific Islander (AIAN/NHPI), Asian, Black, White, Mixed race, and Other. Ethnicity data were collected with caregivers selecting from two options: Hispanic or Non-Hispanic. For highest education level within the household, 29 levels were probed, and collapsed into five groups within the main manuscript: <High School (<HS Diploma; < 13 years), HS/Generalized Education Diploma (GED; ~13 years), Some college (<2 years post HS), Bachelor’s degree (~4 year post HS), Postgraduate degree (>4 years post HS). For household income, 10 levels were collapsed into 3 categories: a) <$50,000; b) $50,000-$99,999; c) ≥$100,000. Caregivers were given the option to answer with “I don’t know” or “refuse to answer”, which were omitted from our analysis.

We incorporated gender identity and sexual health data [8] from KSADS background items on Gender Identity and Sexual Orientation [9] to capture youth that identified as a sexual and/or gender minority. For sexual minorities, we adapted the definitions described by Nagata and colleagues [10]. Participants reported their sexual orientation (“Are you gay or bisexual?” yes, maybe, no, don’t understand the question, decline to answer), where participants who responded “yes” or “maybe” were considered sexual minority adolescents. Participants who reported being transgender or identified with a different gender compared to their sex assigned at birth were considered gender minority participants [8]. Sexual/gender identity data and ED symptom data were matched (e.g., both parent-reported or both youth-reported). Note that for parent reports, sexual and gender identity reports were only available at baseline. For sexual orientation, youth reports at 2-year were used, whereas for gender identity, the closest timepoint (i.e., 3-year) to the youth-reported ED data at 2-year was used.

**SUPPLEMENTARY FIGURES**

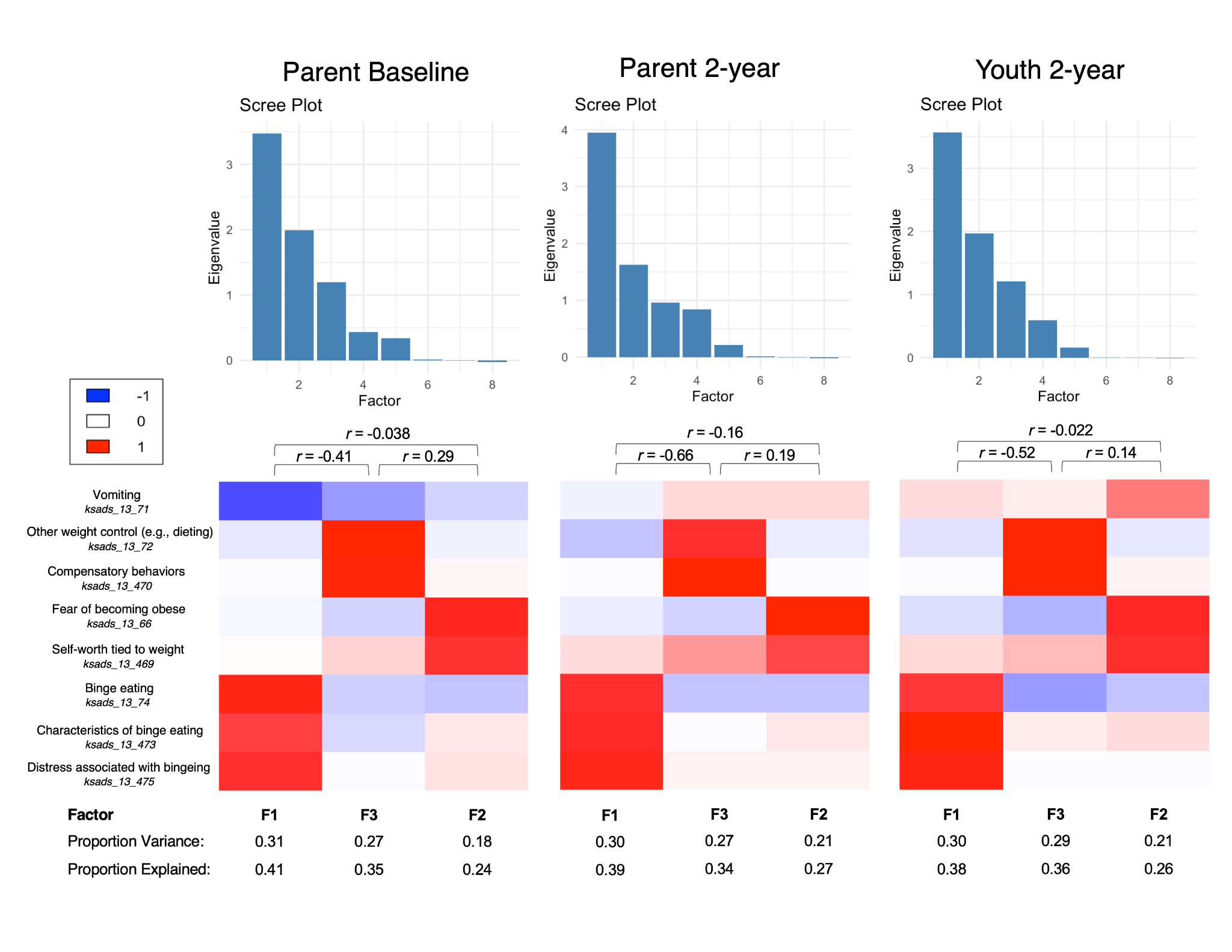

**Supplementary Figure 1**. Three-factor solution from tetrachoric factor analysis of KSADS eating disorder symptoms per timepoint/reporter. Proportion variance listed at the bottom of the figure reflects the overall variance accounted for by a given factor relative to all of the items. Proportion explained reflects the relative amount of variance explained (Proportion variance/sum(proportion variance)). The eight KSADS symptoms used as input into the factor analysis are listed in the bottom left, with the ABCD variable name in italics (i.e., “ksads_13_*”). Correlations between each of the factors are included above each heatmap.

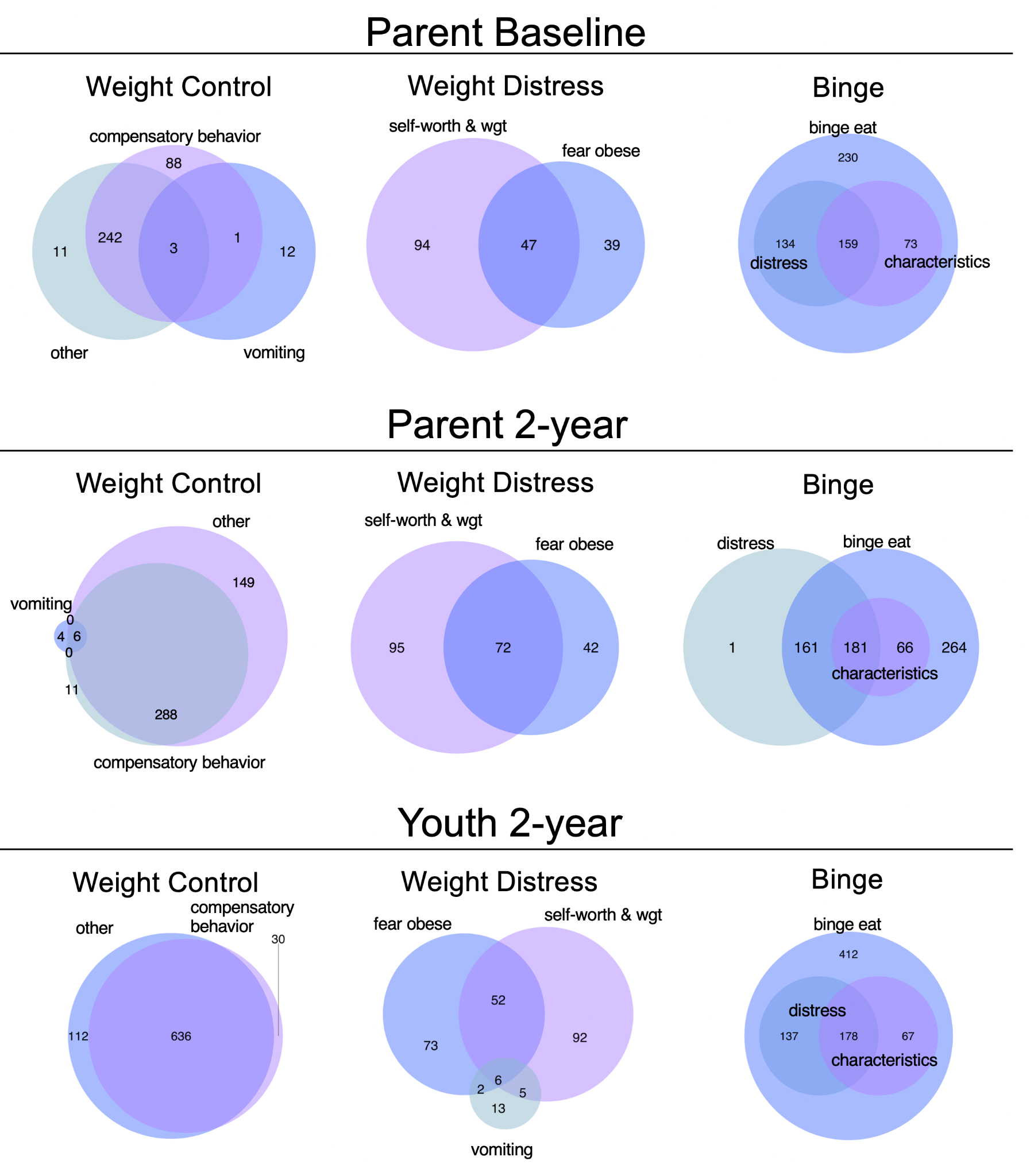

**Supplementary Figure 2**. Breakdown of number of participants endorsing symptoms per cluster, per reporter and timepoint, and overlap in symptom endorsement.
